# Proper Utilization of the National Inpatient Sample Database to Analyze Safety of Cardiovascular Procedures

**DOI:** 10.1101/2025.09.12.25335673

**Authors:** Kathleen Oakes, Jenna Port, Ariel Furer, Lauren Ehrhardt-Humbert, Kevin John, Jennifer Chee, Margaret Infeld, Munther Homoud, Christopher Madias, Fadi Chalhoub, Kevin Heist, Guy Rozen

## Abstract

**Importance:** Analysis of the National Inpatient Sample (NIS) database for the procedural safety of interventions that are primarily outpatient in nature can be inherently biased due to the selective hospitalization of patients who suffer complications during their outpatient procedures.

**Objective:** Using atrial fibrillation (AF) ablation as an example of a procedure typically performed in the outpatient setting, we will test the hypothesis that the population of patients who had their procedure prior to, or on the day of hospital admission (some being outpatient cases, eventually admitted to hospital) will show a significantly higher rate of procedural complications, compared to patients who had a true inpatient ablation, from day 2 of hospital stay and later.

**Design:** This study is a retrospective analysis of hospitalizations in the NIS dataset, including patients aged 18 and older, who underwent an AF ablation between 2016-2022. Patients were divided into two groups based on the day of hospitalization on which the ablation was performed – day 1 of admission or prior (group 1), vs. day 2 of admission or later (group 2). Patient baseline characteristics and procedural complication rates were compared.

**Setting:** Hospitals that performed AF ablations in the United States between January 1, 2016, and December 31, 2022.

**Participants:** A weighted selection of patients aged 18 years and older who underwent an in-patient AF ablation, as identified by the NIS database (N = 47,925).

**Exposures:** Catheter ablation for AF.

**Main Outcome(s) and Measure(s):** In-hospital procedural complications, length of stay and mortality

**Results:** The analysis included 47,925 weighted hospitalizations in patients that underwent AF ablation. Most patients (n= 33,925, 70.8%) underwent catheter ablation (CA) on the day of admission or the days before (group 1). Group 1 was younger, with fewer octogenarians, 6.9% vs. 10.4% (p<0.001), higher proportion of males (60.2% v 58.7%, p=0.003), and had significantly lower rates of comorbid diabetes (24.6% vs. 29.8%), renal disease (12.6% vs. 20.0%), HTN (75.5% vs. 80.1%), heart failure (28.3% vs. 45.8%) and ischemic heart disease (16.0% vs. 20.5%), (p<0.001 for all) compared to Group 2. The rate of complication was higher in group 1 (9.5% v 6.6%, p<0.001). Multivariate analysis showed study group 1 to be an independent predictor of complications (OR 1.54, 95% CI 1.30-1.83, p<0.001).

**Conclusions and Relevance:** Patients undergoing their procedure on the day of admission or prior were younger and had significantly fewer comorbidities, consistent with a population that includes outpatient profile characteristics. Despite the younger and healthier population, these patients suffered from a significantly higher rate of procedural complications. The most plausible explanation to these results is a selection bias, where patients undergoing an outpatient procedure who are at higher risk, or develop complications are admitted as inpatients, thus falsely inflating the complication rate among the patients in the NIS database. The NIS database in its current form, has important limitations when used to assess procedures that are primarily outpatient in nature, requiring careful interpretation of the complication rates arising from such analyses.

## Introduction

The National Inpatient Sample (NIS) database is the largest source of inpatient data publicly available in the US, and provides data on 7 million hospitalizations per year, representing approximately 20% of inpatient hospitalizations and discharges.^1^ Given the large-scale, real-world nature of these data, many analyses use it to assess the safety of cardiovascular procedures in the general population and in various subgroups.

While the NIS is a powerful tool, allowing for real-world, population-level analysis of procedural safety, there is a potential inherent limitation when using this database for procedures that are primarily outpatient in nature. A large meta-analysis of randomized controlled trials (including both inpatients and outpatients) found an overall procedural complication rate of 4.51% for AF ablations.^2^ At the same time, prior studies utilizing the NIS reported much higher complication rates of 6.29%-10.4%.^3,4,5,6,7^

In contemporary practice, most AF ablations are performed in outpatient settings, and patients are discharged same day or observed overnight.^8^ Patients who have higher procedural risk, a more complex procedural course, or develop procedural complications following their outpatient procedure are likely to be admitted to the hospital.

We hypothesized that analyses of the NIS database may overestimate the complication rates of AF ablation, a procedure typically performed in the outpatient setting, because the dataset selectively captures patients admitted with complications or more complex courses. We therefore hypothesize that complication rates in patients who underwent AF ablation prior to, or on the first day of admission will be higher than patients who underwent AF ablations later in the hospital course. Patients who undergo ablation later in the hospital course would represent a less biased sample of inpatient procedures, and their associated complications.

Ablation for AF is only one example of a primarily outpatient procedure whose safety is unlikely accurately estimated by the NIS database, and the scope of this analysis can be expanded to many other procedures in cardiology and in various other fields.

## Methods

Due to restrictions on the data sharing in the Healthcare Cost and Utilization Project Data Use Agreement, NIS data, statistical methods, study material, and analytical tools used for this study will not be made available to other researchers. However, the NIS database is publicly available for purchase and the detailed materials and methods described below will make it possible for anyone to replicate this study and reproduce our results.

### Data Source

Data was obtained from the NIS, a nationwide database developed for the Healthcare Cost and Utilization Project and the largest publicly available collection of all-payer inpatient data for hospitalizations in the United States (US).^1^ Unweighted, it provides data on 7 million hospitalizations per year, amounting to approximately 20% of inpatient discharges from US hospitals (47 states in total). The data can be used to calculate national estimates by using the sampling weights provided by the NIS. The database contains both patient-level and hospital-level factors; this includes patient demographics, primary and secondary hospitalization diagnoses, procedure codes and timing, length of stay, mortality, as well as hospital bed size, region, and teaching status. For this study, we obtained data for the years 2016 to 2022. As the NIS datasets only include deidentified data, the study is deemed exempt from institutional review by the Human Research Committee. Further information regarding the NIS database is provided in the supplemental material (Data S1).

### Study Population and Variables

The NIS dataset includes basic demographic data (age, sex, race) which we collected to assess differences in the baseline characteristics for our populations of interest. For our study period, the NIS reported diagnosis and procedure codes using the *International Classification of Diseases, Tenth Revision, Clinical Modification* (*ICD-10-CM*). Our target population was patients, aged 18 years or older, who had a diagnosis code of AF (*ICD-10-CM* I480, I4891, I482, I4819, I481, I489, I480, I4811, I4821, I48, I4820) as well as a procedure code for a CA (*ICD-10-CM* 025S3ZZ, 025T3ZZ, 02573ZZ, 02583ZZ). To identify only those with AF, we excluded patients having a diagnosis of atrial flutter, pre-excitation syndromes, premature beats, paroxysmal tachycardia as well as those who have the codes for the presence of a pacemaker or defibrillator. To avoid the inclusion of patients who underwent a procedure for ablation of the atrioventricular junction, we excluded patients with diagnostic or procedural codes for defibrillator and pacemaker placements or adjustments. Additionally, we excluded any open surgical procedure. This methodology has been used in several studies to identify patients undergoing AF catheter ablation from NIS databases.^3,4,5,6,7^

We classified patients into groups based on the day of their procedure, which is documented in the NIS database. The NIS also includes procedures that occurred up to 4 days prior to hospitalization, denoted by a negative integer (-4 = procedure 4 days prior to admission). The NIS database denotes the first day of admission as “0”, however throughout this paper we refer to the first day of admission as day 1 for simplicity. We looked for the AF ablation procedure day and classified the patients into two groups. We assumed that some of the cases where ablation was performed on day 1 of the hospitalization, likely represented patients who were admitted following an outpatient procedure, as patients admitted to the hospital would be very unlikely to undergo an “urgent” AF ablation on the first day of admission. We included all patients with procedures on day 1 and earlier in group 1, while all patients with procedures on day 2 of the admission and later were included in group 2. Patients who did not have a procedure day listed were excluded from the final analysis (n=186).

The specific *ICD-10-CM* inclusion and exclusion codes used are listed in (supplemental) Table S1.

### Study Outcomes

Our primary study outcomes were the known potential complications for an AF catheter ablation, as described in prior AF ablation studies.^3,4,5,6,7^ The complications analyzed in this paper included pericardial (hemopericardium, tamponade, need for pericardiocentesis, pericarditis), cardiac (cardiac arrest, arrythmia, myocardial infarction, congestive heart failure), pulmonary (pneumothorax, hemothorax, respiratory failure, diaphragm paralysis, other respiratory compromise), intra-op or post-procedural hemorrhage or hematoma, vascular (complication requiring surgical repair, accidental puncture/laceration, arteriovenous fistula), infection (including bacteremia and sepsis), and neurologic (cerebrovascular accident, transient ischemic attack) complications. These codes were selected based on a review of pertinent literature and matched with other studies as closely as possible to maintain consistency.^3,4,5,6,7^ Please see Table S3 for the complete list of complication *ICD-10-CM* codes utilized.

### Statistical Analysis

All statistical analysis was performed on weighted data, which was obtained by applying the weights provided by the NIS to the unweighted discharge data in the database. Descriptive statistics were presented as numbers, with percentages for the categorical variables. Means and standard deviations were reported for continuous variables. A Pearson χ^2^ was used to calculate *p*-values for categorical variables, and *t*-tests were used for continuous variables. Multivariate logistic regression was used to identify independent predictors of complications. Variables included in the regression models were the following comorbidities: age, diabetes, renal disease, hypertension, cardiomyopathy, history of stroke, ischemic heart disease, obesity, and obstructive sleep apnea (OSA).

Data analysis and visualization were performed using Python 3.12.7 in Jupyter Notebook 7.2.2. The pandas 2.2.1 library was used for data handling, while NumPy 1.26.4, SciPy 1.12.0, and statsmodels.api 0.14.2 were used for statistical analysis. Data visualization was performed with matplotlib 3.8.3. A *p*-value of < 0.05 was considered statistically significant.

## Results

### Baseline Characteristics

A total of 47,925 weighted hospitalizations were included in the final analysis. Group 1 made up most hospitalizations (n = 33,925, 70.8%). The median day of procedure in Group 1 was 1.0 (IQR 1.0-1.0) and the median day of the procedure was 4.0 (IQR 2.0 – 6.0) in Group 2. The median age was 67.0 (IQR 59.0 – 72.0) in Group 1 and 68.0 (IQR 60.0 – 74.0) in Group 2. Overall, Group 1 had fewer octogenarians, 6.9% vs. 10.4% (p<0.001) and a higher proportion of males (60.2% v 58.7%, p=0.003). Furthermore, patients in Group 1 had significantly lower rates of comorbid diabetes (24.6% vs. 29.8%), renal disease (12.6% vs. 20.0%), HTN (75.5% vs. 80.1%), heart failure (28.3% vs. 45.8%) and ischemic heart disease (16.0% vs. 20.5%), (p<0.001 for all) compared to Group 2. Baseline characteristics are detailed in Table 1.

**Table 1:** Baseline Characteristics of the Study Population.

| **Table 1:** Baseline Characteristics of the Study Population | Overall | Day 1 or Prior | Day 2 or Later | p-value |
| --- | --- | --- | --- | --- |
| **Unweighted count, n** | 9585 | 6785 | 2800 |  |
| **Weighted count, n** | 47925 | 33925 | 14000 |  |
| **Age** |  |  |  |  |
| 18-34 y | 0.8% (385) | 0.8% (280) | 0.8% (105) | 0.433 |
| 35-49 y | 5.9% (2834) | 6.0% (2034) | 5.7% (799) | 0.239 |
| 50-64 y | 32.0% (15344) | 32.7% (11089) | 30.4% (4255) | < 0.001 |
| 65-79 y | 53.3% (25549) | 53.6% (18170) | 52.7% (7379) | 0.094 |
| >80 y | 7.9% (3809) | 6.9% (2350) | 10.4% (1459) | < 0.001 |
| **Sex** |  |  |  |  |
| Male | 59.7% (28634) | 60.2% (20414) | 58.7% (8219) | 0.003 |
| Female | 40.3% (19290) | 39.8% (13510) | 41.3% (5780) | 0.003 |
| **Race** |  |  |  |  |
| White | 81.9% (39229) | 82.4% (27954) | 80.5% (11274) | < 0.001 |
| Black | 5.1% (2454) | 4.5% (1534) | 6.6% (919) | < 0.001 |
| Hispanic | 5.8% (2769) | 5.5% (1864) | 6.5% (905) | < 0.001 |
| Asian or Pacific Islander | 1.8% (850) | 2.0% (665) | 1.3% (184) | < 0.001 |
| Native American | 0.3% (145) | 0.3% (100) | 0.3% (44) | 0.695 |
| Other | 3.2% (1520) | 3.3% (1135) | 2.7% (384) | < 0.001 |
| **Payment type** |  |  |  |  |
| Medicare | 58.5% (28049) | 57.1% (19354) | 62.1% (8694) | < 0.001 |
| Medicaid | 5.4% (2574) | 5.0% (1699) | 6.2% (874) | < 0.001 |
| Private Insurance | 32.4% (15519) | 34.5% (11704) | 27.3% (3815) | < 0.001 |
| Self-pay | 1.0% (499) | 0.8% (274) | 1.6% (225) | < 0.001 |
| No Charge | 0.1% (59) | 0.1% (25) | 0.2% (34) | < 0.001 |
| Other | 2.5% (1200) | 2.5% (844) | 2.5% (355) | 0.799 |
| **Hospital Region** |  |  |  |  |
| Northeast | 19.6% (9404) | 21.3% (7209) | 15.7% (2195) | < 0.001 |
| Midwest | 11.9% (5695) | 12.4% (4215) | 10.6% (1480) | < 0.001 |
| South | 30.7% (14709) | 28.6% (9709) | 35.7% (4999) | < 0.001 |
| West | 9.7% (4644) | 9.9% (3369) | 9.1% (1274) | 0.006 |
| **Comorbidities** |  |  |  |  |
| Diabetes | 26.1% (12529) | 24.6% (8354) | 29.8% (4174) | < 0.001 |
| Renal | 14.8% (7080) | 12.6% (4275) | 20.0% (2804) | < 0.001 |
| Hypertension | 76.9% (36834) | 75.5% (25619) | 80.1% (11214) | < 0.001 |
| Heart Failure | 33.4% (16009) | 28.3% (9604) | 45.8% (6404) | < 0.001 |
| CVA | 8.7% (4189) | 8.9% (3019) | 8.4% (1169) | 0.057 |
| Ischemic heart disease | 17.3% (8299) | 16.0% (5429) | 20.5% (2869) | < 0.001 |
| Obesity | 30.3% (14524) | 29.8% (10119) | 31.5% (4404) | < 0.001 |
| OSA | 25.4% (12149) | 26.0% (8809) | 23.9% (3340) | < 0.001 |

*P values were calculated with chi-squared tests to analyze the difference in frequency of the specified characteristics among patients with CA procedures done on day 0 of admission or earlier versus day 1 of admission or later. CA = catheter ablation; AF = atrial fibrillation; CVA = cerebrovascular accident*

### Rates of In-Hospital Complications

Despite Group 1 being younger and healthier, these patients had a significantly higher rate of overall complications, 8.8% v 6.5% (p<0.001), consistent with selective capture of outpatients who developed complications or required closer monitoring. Complication rates were higher in Group 1 across almost all categories, including pericardial (4.4% v 2.2%), cardiac (1.0% v 0.7%), hemorrhage/hematoma (1.6% v 1.2%), and vascular (1.9% v 0.8%, (p<0.001 for all). Conversely, Group 1 had a significantly lower rate of infectious complications (0.5% v 0.9%, p<0.001). Complication rates by category are detailed in Table 2.

**Table 2:** Complications Overall and by Category Between Procedure Day Groups.

Complication Rates by Procedure Day
|  | Overall | Day 1 or Prior | Day 2 or Later | p-value |
| --- | --- | --- | --- | --- |
| **Complications** |  |  |  |  |
| **Cardiac** | 1.0% (465) | 1.0% (354) | 0.9% (120) | 0.105 |
| **Hemorrhage/Hematoma** | 1.6% (755) | 1.8% (600) | 1.1% (155) | <0.001 |
| **Infection** | 0.6% (300) | 0.5% (160) | 1.0% (140) | <0.001 |
| **Neurologic** | 0.4% (175) | 0.5% (155) | 0.1% (20) | <0.001 |
| **Pericardial** | 3.8% (1815) | 4.5% (1530) | 2.0% (285) | <0.001 |
| **Respiratory** | 1.6% (745) | 1.6% (545) | 1.4% (200) | 0.152 |
| **Vascular** | 1.8% (860) | 2.0% (685) | 1.3% (175) | <0.001 |
| **Any complication** | 8.6% (4145) | 9.5% (3225) | 6.6% (920) | <0.001 |

*P values were calculated with chi-squared tests to analyze the difference in frequency of the specified complication rates among patients with CA procedures done on day 0 of admission or earlier versus day 1 of admission or later. Complications were defined as at least one complication in each respective category.*

**Table 3:**
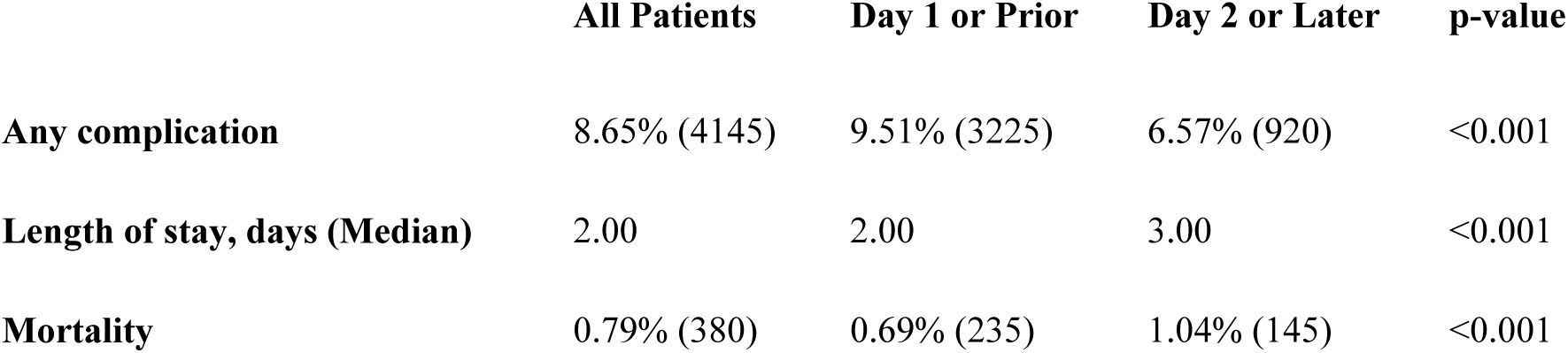
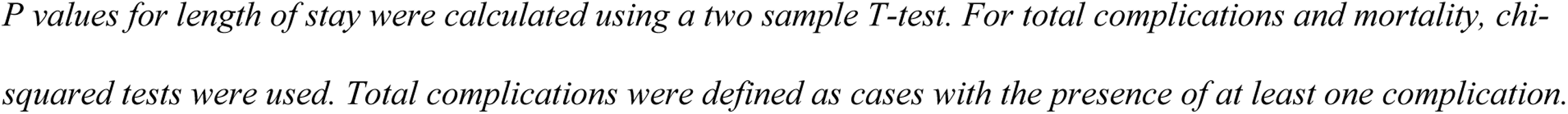
Complications, Length of Stay, and Mortality Differences Between Procedure Day Groups.

|  | All Patients | Day 1 or Prior | Day 2 or Later | p-value |
| --- | --- | --- | --- | --- |
| **Any complication** | 8.65% (4145) | 9.51% (3225) | 6.57% (920) | <0.001 |
| **Length of stay, days (Median)** | 2.00 | 2.00 | 3.00 | <0.001 |
| **Mortality** | 0.79% (380) | 0.69% (235) | 1.04% (145) | <0.001 |

*P values for length of stay were calculated using a two sample T-test. For total complications and mortality, chi-squared tests were used. Total complications were defined as cases with the presence of at least one complication.*

**Table 4:** Multivariable Analysis of Predictors of Complications in Patients Undergoing CA for AF.

**Multivariate Analysis**
|  | Odds Ratio | 95% Confidence Interval | p-value |
| --- | --- | --- | --- |
| **Sex** |  |  |  |
| Male |  | reference |  |
| Female | 1.23 | 1.07 - 1.44 | 0.005 |
| **Race** |  |  |  |
| White |  | reference |  |
| Non-White | 0.82 | 0.67 - 1.01 | 0.069 |
| **Age** |  |  |  |
| 18-34 y |  | reference |  |
| 35-49 y | 0.90 | 0.34 – 2.38 | 0.836 |
| 50-64 y | 0.99 | 0.39 – 2.51 | 0.986 |
| 65-79 y | 1.07 | 0.42 - 2.70 | 0.892 |
| >80 y | 1.01 | 0.39 - 2.65 | 0.976 |
| **Comorbidities** |  |  |  |
| Procedure Day 1 or Prior | 1.54 | 1.30 - 1.83 | <0.001 |
| Diabetes | 0.99 | 0.84 - 1.18 | 0.947 |
| Heart Failure | 1.03 | 0.88 - 1.21 | 0.734 |
| Ischemic heart disease | 0.88 | 0.73 - 1.08 | 0.223 |
| OSA | 0.96 | 0.81 - 1.14 | 0.667 |
| Obesity | 1.39 | 1.19 - 1.63 | <0.001 |
| Renal | 1.34 | 1.10 - 1.64 | 0.004 |
| Hypertension | 1.07 | 0.89 - 1.29 | 0.470 |
| CVA | 1.56 | 1.25 - 1.94 | <0.001 |

CVA - cerebrovascular accident; OSA - obstructive sleep apnea

### Length of Stay and Mortality

Length of stay was shorter in Group 1, with an average of 3.0 (median 2.0 IQR 1.0 – 3.0) days versus 5.44 (median 4.0 IQR 2.0 – 7.0) days in Group 2, p<0.001. Mortality was significantly lower in Group 1 (0.69% v 1.04%, p<0.001).

### Predictors of Complications

Multivariable analysis (including age, sex, race and different comorbidities) showed renal failure (OR 1.28, 95% CI 1.04 – 1.56, p=0.019), obesity (OR 1.40, 95% CI 1.19 – 1.64, p<0.001), female sex (OR 1.24, 95% CI 1.07 – 1.44, p=0.005) to be an independent predictor of complications. Importantly, study group 1 (procedures, done prior or on day 1 of the admission), was found to be an independent predictor of complications, (OR 1.54, 95% CI 1.30-1.83, p<0.001), (Figure 3).

**Figure 1:**
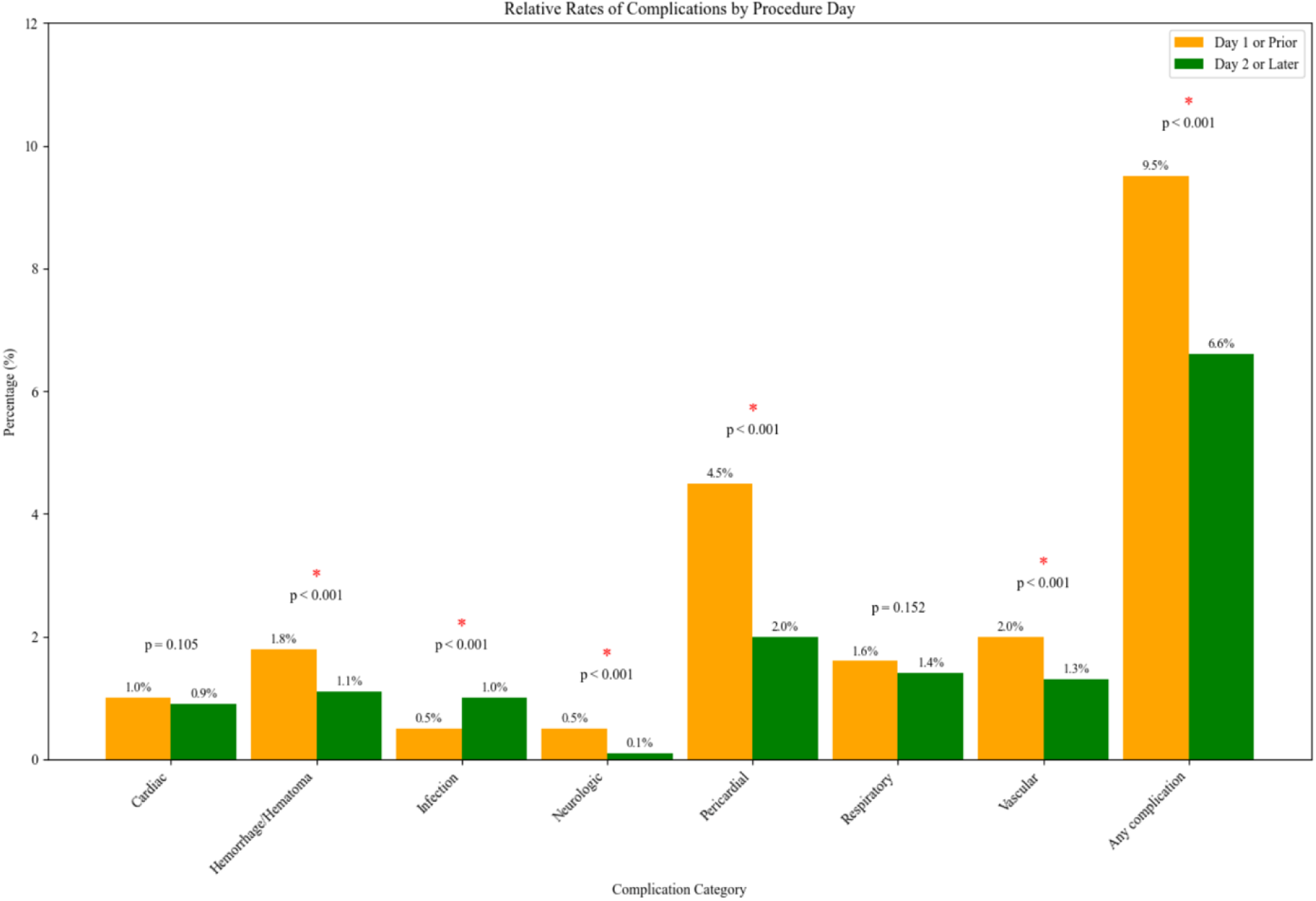
Relative Rates of Complications by Procedure Day Group.

**Figure 2:**
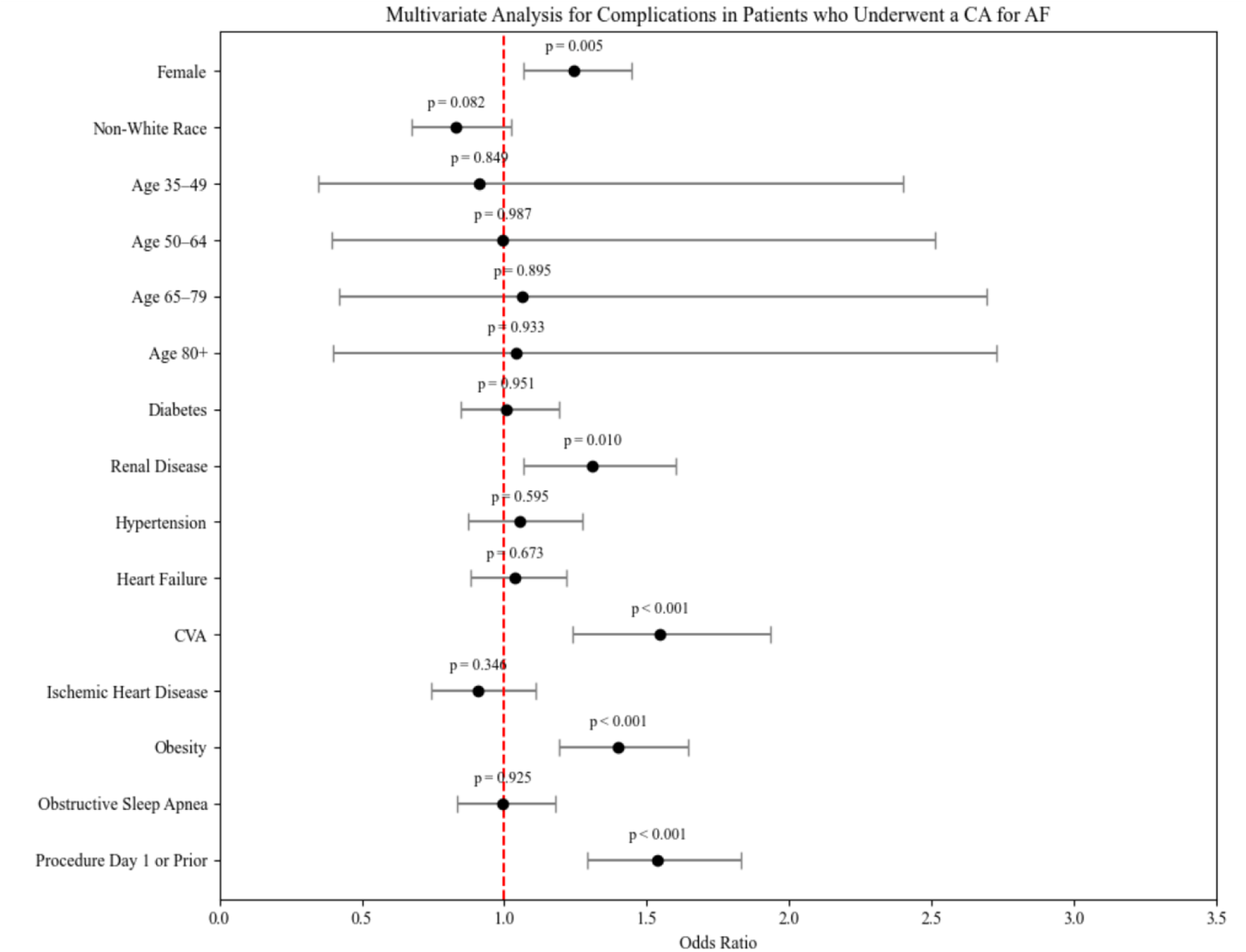
Multivariable Analysis of Complications.

**Figure 3:**
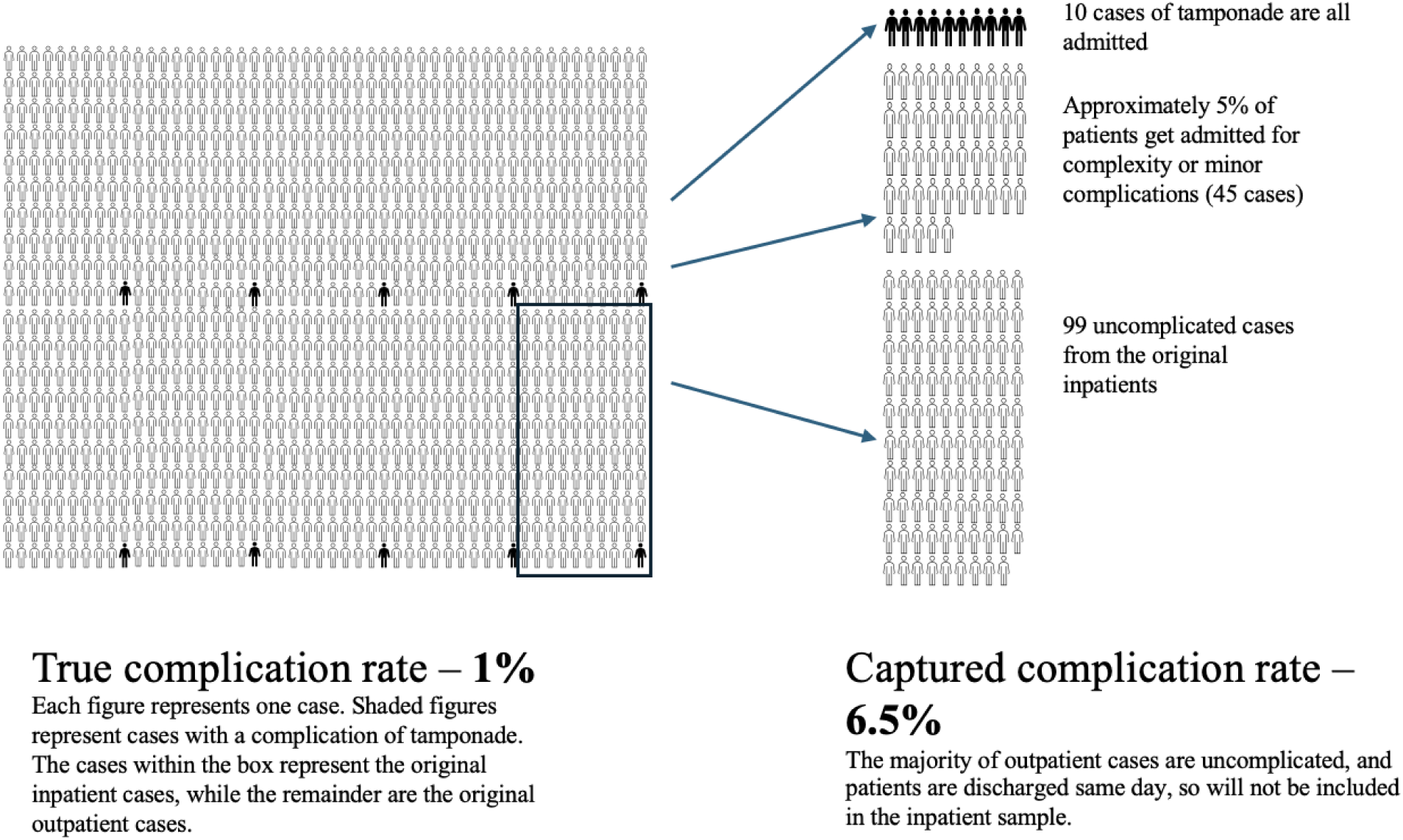
Visual Representation of Selection Bias in the NIS.

## Discussion

Using data from the NIS, the largest inpatient database in the United States, we analyzed the baseline characteristics and rates of complications of CA for AF, in patients who had their ablation procedure before or on day 1 of their hospitalization, compared to day 2 of the hospitalization and later. Patients who underwent their ablation on the day of admission or prior were younger and had significantly fewer comorbidities, consistent with a population that includes outpatient profile characteristics. Despite the younger and healthier population, these patients suffered from a significantly higher rate of procedural complications.

We believe the most plausible explanation for these results is a selection bias. Patients undergoing outpatient procedures who are at higher risk or subsequently develop complications are likely to be admitted as inpatients, thus falsely inflating the actual inpatient procedural complication rate among the patients in the NIS database. As an example, if 1000 patients are undergoing a procedure with a known risk of 1% for tamponade and 90% of these cases are done on an outpatient basis, and 100 as inpatients, there will be 9 cases of tamponade out of 900 outpatient cases and 1 case of tamponade out of 100 inpatients. Out of the patients that had their procedure done electively, all patients with tamponade would be admitted, along with a certain percentage (we can say 5% for this example) of 900 outpatients (45 patients in our example) with complex procedural course, or minor complaints. This will result in total of 154 patients that are eventually listed as inpatients, out of which 10 have had a tamponade. Analyzing the data on the inpatients only will result in tamponade rate of 10 out of 154 cases, which is clearly inaccurate, due to the selection bias, including a disproportionate number of the outpatients with complications that are eventually being admitted. This example is illustrated visually in Figure 3.

Additionally, other studies comparing AF ablation between inpatient and outpatient populations have demonstrated higher rates of procedural complications in inpatient groups, likely due to higher rates of comorbidities among these patients.^8^ This further suggests the presence of a selection bias within the NIS.

The baseline characteristics of patients who had their procedures on day 1 or earlier (Group 1) versus during hospitalization (Group 2) were significantly different. Group 1 was overall younger and healthier, with significantly lower rates of comorbid diabetes, CKD, HTN, CAD, and heart failure. This was expected, as healthier patients are more likely to be planned for an outpatient procedure, while sicker patients may have been admitted to the hospital for closer monitoring or optimization prior to undergoing a procedure. Additionally, it is possible that older and sicker patients admitted to the hospital for uncontrolled AF, or heart failure with AF, are likely to have failed other treatment strategies, thus necessitating a more aggressive treatment approach.

Despite Group 1 being younger and healthier, and having a higher male/female ratio, they had significantly higher overall complication rates. This fits with our hypothesis that the utilization of the NIS database to analyze complications of a procedure that is mostly outpatient in nature presents a significant inherent bias. Many patients who undergo an uncomplicated procedure can be discharged the same day of the procedure or the day after (“bedded outpatient” cases) and are not included in the NIS database.

Patients undergoing AF ablation who are subsequently admitted following the procedure, are more likely to be sicker patients requiring closer monitoring, or those that suffered an acute complication. The overall complication rate in the NIS database for procedures that are mainly outpatient in nature would thus be falsely elevated. Among the categories of complications, pericardial, cardiac, hemorrhage/hematoma, and vascular complications were all significantly higher in Group 1. These complications would warrant switching the patient to inpatient status, although the case could have been originally planned to be done in outpatient setting. Only infectious complications were significantly higher in Group 2, which is not surprising given that the older and sicker patients in group 2 had longer hospital stays, putting them at risk for nosocomial infections.

Length of stay was significantly shorter in Group 1. The likely explanation here is that while Group 1 had higher rates of complications, this rate is still low and there are many patients who may have been admitted for observation after a procedure but who did not suffer a significant complication that was captured by our analysis. These might include patients admitted for groin oozing, monitoring for small pericardial effusion or chest pain due to pericardial irritation, that would be potentially warranting 1 additional day of observation before discharge. Even when these patients did suffer a complication, the fact that they were younger and healthier and that most of these complications, including the more dangerous ones such as pericardial effusion that required pericardiocentesis, would in many cases prolong these hospitalizations by another couple of days, and the patients quickly recovered. Further, group 2 also represented patients who are admitted for symptomatic, difficult to control atrial fibrillation, many with heart failure symptoms, who failed more conservative measures (i.e. cardioversion, antiarrhythmics), so the decision to ablate as an inpatient could have met scheduling difficulties prolonging their stay to wait for the procedure that may have been performed a few days later.

Mortality was significantly lower in Group 1. This again is likely reflective of the fact that this is a younger and healthier group. Although they have higher rates of complications, they would be more likely to survive a complication than an elderly and frail patient.

To our knowledge, this is the first study to identify an important inherent bias in analysis of complication rates for cardiovascular interventions in the NIS database that are generally performed on an outpatient basis. The results of our analysis suggest that “procedure day” should be accounted for in future studies aiming to analyze true in-patient procedure complications, and a more accurate analysis should likely be limited to patients who had their procedures performed on day 2 or later.

### Limitations

The NIS database is a retrospective database and is susceptible to coding errors. There are many institutions represented in the NIS database which may have different reporting and coding procedures. Our study is an observational and noncontrolled cohort study, so we cannot make direct conclusions on causality. There is no standardized approach in the literature for which ICD-10 codes should be used to represent complications. While we attempted to synthesize the methods used by other studies, there is no way to include all possible codes that represent complications, nor ascertain that these codes are truly reflective of complications and not unrelated to the procedure of interest.

## Conclusion

Patients undergoing their AF ablation procedure on the day of admission or prior were younger and had significantly fewer comorbidities, consistent with population that includes outpatient profile characteristics. Despite the younger and healthier population, these patients suffered from a significantly higher rate of procedural complications. The most plausible explanation to these results is a selection bias, where patients undergoing an outpatient procedure who are at higher risk, or develop complications are admitted as inpatients, thus falsely inflating the complication rate among the patients in the NIS database. We conclude that the NIS database cannot be used, as is, to accurately assess the “real-world” risk of mainly outpatient in nature procedures. The NIS database in its current form, has important limitations when used to assess procedures that are primarily outpatient in nature, requiring careful interpretation of the complication rates arising from such analyses. Future analyses of “inpatient procedures”, for these types of procedures should consider concentrating on patients that had their procedures performed on day 2 or later of the hospitalization.

## Data Availability

Due to restrictions on the data sharing in the Healthcare Cost and Utilization Project Data Use Agreement, NIS data, statistical methods, study material, and analytical tools used for this study will not be made available to other researchers. However, the NIS database is publicly available for purchase and the detailed materials and methods described in our analysis will make it possible for anyone to replicate this study and reproduce our results.

https://hcup-us.ahrq.gov/db/nation/nis/nisdbdocumentation.jsp

## Acknowledgments

N/A

